# Bibliometric analysis of research areas, publication hierarchy and gender authorship in German university cardiology

**DOI:** 10.64898/2025.12.17.25342464

**Authors:** Eike Sebastian Debus, Markus Geissen, Stefan Blankenberg, Yskert von Kodolitsch, Reinhart T. Grundmann

## Abstract

This bibliometric study focuses on the publication activity of senior staff, consisting of chief physicians, senior physicians, department heads, and division heads at German university hospitals specializing in cardiology. The aim is to identify topics, focal points, study types, and impact factors (IFs) achieved. No studies on this topic have been conducted to date. However, Millenaar et al. [1] provided an overview of the cardiovascular research landscape in Germany, highlighting publication activities by federal state, the gender distribution among first authors, and the most important areas of cardiological research. In a subsequent global scientometric analysis, Millenaar et al. [2] examined gender differences in authorship in cardiovascular research between 2010 and 2019. They found that the number of publications in cardiovascular research has increased over the last ten years, particularly among female authors. However, a detailed analysis of the associated IFs revealed that articles by female authors achieved a lower average IF compared to their male counterparts. Similarly, publications by female authors were cited less frequently. The aim of this study was to provide an overview of the publishing activities of senior staff in the cardiology departments of all 39 university hospitals. The hierarchy, academic rank, and gender of the authors were taken into account. The aim was to determine which specialist areas and topics are emphasized in cardiology publications and which impact factors were assigned to the respective areas. Furthermore, the composition of the hierarchical levels according to gender and academic rank of the management staff and the respective publication performance (number; impact factor) were determined. For comparison purposes, the methodological approach was adapted to previous bibliometric analyses in other medical fields such as visceral surgery [3], trauma surgery [4], and oral and maxillofacial surgery [5]. The aim was to investigate whether the pronounced gender-specific differences in publication output and academic leadership roles observed in these fields can also be observed in cardiology. This work can serve as a basis and reference for the effectiveness and implementation of measures to promote women.

## Materials and Methods

For the analysis, the publication output of the current senior staff (director, deputy director, senior staff, department heads, senior physicians) from 39 German university cardiology departments was recorded. Senior staff from research institutions were included if they were assigned to the cardiology faculty on the official website. For academic classification, a distinction was made between “university professor”; “assistant professor”; “doctor” and “no academic title.” The composition of the staff, position, and academic degree were taken from the departments’ websites. The cut-off date was September 1, 2021. No distinction was made according to the type of doctoral title (Dr. med; Dr. rer. nat.; Dr. phil. nat., etc.) was not made. For the database search in PubMed, all publications by the listed employees (only first and last authors; co-authors were not included) over a period of ten years (September 1, 2011, to August 30, 2021) were recorded. To be included in the evaluation, publications had to meet the following criteria: Listing in PubMed (Medline) as an original article with abstract (no comments or responses). The full name of each employee was used for the search. Since joint first authorship (last authorship) is not apparent in PubMed, publications were assigned to the first author listed (or the last author listed). Broderick 2019 showed that for publications after 2007, no statistically significant gender bias in the order of first authors is to be expected [26]. In the case of double names, an additional PubMed search was performed using the individual surnames in combination with the first name. Non-medical literature was checked for plausibility of the topic (or subject area) of the publication and the author’s place of work in order to exclude publications by authors with the same name. Gender was assigned based on first names and photos on the website.

The publications were categorized based on the following criteria: author’s name; academic title; first/last author; journal; impact factor; field (angiology, noninvasive diagnostics, intensive care medicine, interventional cardiology, cardiology, neurology, palliative medicine, pulmonology); topic (angiography/cardiac catheterization; education and training; biomarkers; diagnostics; device evaluation; gender-specific; complications; costs; medication; outcome; pathogenesis; prevention; sports cardiology/exercise; telemedicine; therapy; survey); Focus; Study type (observational study; case report; human research; basic research; laboratory study; guidelines; meta-analysis; prospective study; randomized study; retrospective study; review; overview article); DOI. The subcategories for specialty, topic, and study type were defined prior to data analysis. The “focus” list was continuously updated during the analysis and only after indexing into higher-level thematic categories (heart rhythm; heart failure; coronary heart disease; heart valves; vascular system; invasive therapy; defined comorbidities; other comorbidities; laboratory/technology/documentation). The 5-year impact factor from 2016 was used for the journal’s impact factor (jcr.clarivate.com). If a journal did not have a 5-year impact factor for 2016, the value from 2020 was used instead. Journals with an IF = 0 were counted as 0. Publications were always attributed to the department where the respective author was employed on the cut-off date.

Before analyzing publication performance by clinic, clinic IF, study type, and journal analysis, all duplicates (1,123 in total) were removed. This left 7,156 publications in 876 journals. A duplicate was considered to exist if both the first and last authors of a publication were employed in the same department on the reference date. All publications (n = 8,279) were included in the analysis of publication performance by hierarchical level, gender, or academic title.

## Results

### Subject

The cardiology departments of German university hospitals have published articles in a wide range of specialist areas (Table 1). As expected, cardiology (51.52% of publications) and interventional cardiology (32.29%) topics were the most common subjects of the publications. These were followed by “Imaging diagnostic methods” (13.25%), angiology (peripheral) with 4.85%, and “intensive care / emergency medicine” (2%). Pneumology (1%) and neurology (0.45%) accounted for only a small proportion of the publications. It should be noted that publications on neurological topics achieved the highest IF per publication at 5.89, followed by angiology (peripheral) at 5.69 and cardiology at 4.89. The number of university hospitals that have published articles on the specific subjects, as well as further details, are listed in Table 1.

**Table 1:** Publication activity by subject in 39 cardiology departments based on the total number of publications (n=7156). All double authorships were excluded from this table. c-IF = cumulative impact factors

| Publication activity by subject |  |  |  |  |  |  |
| --- | --- | --- | --- | --- | --- | --- |
| subject | pub [n] | publ [%] | c-IF | c-IF [%] | IF pro pub | Universities |
| Cardiology | 3230 | 45,14 | 15795,46 | 51,52 | 4,89 | 38 |
| Interventional cardiology | 2311 | 32,29 | 8318,57 | 27,13 | 3,60 | 38 |
| Imaging diagnostic methods | 948 | 13,25 | 3603,85 | 11,75 | 3,80 | 37 |
| Angiology (periphery) | 347 | 4,85 | 1972,79 | 6,43 | 5,69 | 26 |
| Intensive care/emergency medicine | 217 | 3,03 | 631,08 | 2,06 | 2,91 | 32 |
| Pneumology | 71 | 0,99 | 151,62 | 0,49 | 2,14 | 7 |
| Neurology | 32 | 0,45 | 188,43 | 0,61 | 5,89 | 12 |
| Total | 7156 | 100,00 | 30661,82 | 100,00 |  |  |

### Topics

The most important publication topics and the cumulative impact factors (c-IF) achieved are listed in Table 2. “Outcome” was the most frequently covered topic with 20.1% of publications and achieved the highest c-IF (6,583.9), followed by ‘Diagnostics’ (16%, 4,163.6 c-IF) and “Therapy” (15.6%, 4,041.5 c-IF). The highest average impact factor per individual publication was achieved in the areas of “Pathogenesis” (average IF 5.8) and ‘Biomarkers’ (average IF 5.2), while the lowest values were recorded in the areas of “Education and Training” (IF 2.2) and “Device Testing” (IF 2.9).

**Table 2:** Publication activity by topic in 39 cardiology departments based on the total number of publications (n=7156). All double authorships were excluded from this table. c-IF = cumulative impact factors

| Publication activity by topic |  |  |  |  |  |
| --- | --- | --- | --- | --- | --- |
| topic | publikation [n] | c-IF | IF/publikation | publikation [%] | c-IF [%] |
| Outcome | 1438 | 6583,9 | 4,6 | 20,1 | 21,5 |
| Diagnostics | 1147 | 4163,6 | 3,6 | 16,0 | 13,6 |
| Therapy | 1114 | 4041,5 | 3,6 | 15,6 | 13,2 |
| Complications | 1069 | 4493,3 | 4,2 | 14,9 | 14,7 |
| Biomarkers | 722 | 3743,5 | 5,2 | 10,1 | 12,2 |
| Pathogenesis | 653 | 3784,9 | 5,8 | 9,1 | 12,3 |
| Medication | 546 | 2275,7 | 4,2 | 7,6 | 7,4 |
| Gender-specific | 78 | 287,9 | 3,7 | 1,1 | 0,9 |
| Training and Education | 72 | 157,9 | 2,2 | 1,0 | 0,5 |
| Prevention | 72 | 245,6 | 3,4 | 1,0 | 0,8 |
| Survey | 51 | 180,1 | 3,5 | 0,7 | 0,6 |
| Sports cardiology/exercise | 50 | 205,7 | 4,1 | 0,7 | 0,7 |
| Device evaluation | 42 | 123,1 | 2,9 | 0,6 | 0,4 |
| Angiography/cardiac catheterization | 38 | 136,1 | 3,6 | 0,5 | 0,4 |
| Telemedicine | 38 | 152,4 | 4,0 | 0,5 | 0,5 |
| Costs | 26 | 86,6 | 3,3 | 0,4 | 0,3 |
| Summe | 7156 | 30661,8 |  | 100,0 | 100,0 |

### Publication Focus

An important aspect of the analysis is the classification of publications in relation to their focus. Which clinical pictures are the focus of clinical consideration? Articles on coronary heart disease accounted for the largest share of publications at 19.9%. These also achieved the highest value in terms of cumulative impact factor at 22.5% (Table 3), followed by studies on cardiac arrhythmias (14.7% of all publications / 12.3% of the c-IF) and heart failure (13.6% of publications / 15.3% of the c-IF). The focus on coronary heart disease also achieved one of the highest IF per individual publication (4.8), together with heart failure and “other comorbidities,” which included ambiguous symptoms/clinical pictures such as hypoxia, muscle wasting, obesity, environmental influences, and sepsis, while publications on cardiac arrhythmias achieved an average of 3.6 IF/publication and those on invasive therapies achieved 3.5 IF/publication. Articles on heart failure (13.6%), atrium (9.98%), and heart attack (7.7%) were published most frequently. Publications on TAVI/TAVR represented (4.79%), arrhythmias (4.72%), and heart valves (4.72%) over the entire observation period. CAD, cardiovascular aspects, and articles on pacemakers were also frequently represented, each accounting for slightly more than 3%. Also worth mentioning is the relatively high proportion of literature on more peripheral/vascular medicine topics (arteriosclerosis 3.7%; blood pressure 3%; vascular blood vessels 1.37%; PAOD 1.16%), which reflects the variable interdisciplinary orientation of some university hospitals in Germany, where cardiology, angiology, vascular surgery, and pneumology are combined in various ways within a single hospital.

**Table 3:** Number and IF of research foci of all publications. All double authorships were excluded from this table. It is based on the total number of publications (n=7156); c-IF = cumulative impact factors

| Publication activity by focus |  |  |  |  |  |
| --- | --- | --- | --- | --- | --- |
| focus | publikation [n] | c-IF | IF/publikation | publikation [%] | c-IF [%] |
| Coronary heart disease | 1427 | 6900,5 | 4,8 | 19,9 | 22,5 |
| Heart rhythm | 1052 | 3786,0 | 3,6 | 14,7 | 12,3 |
| Heart failure | 973 | 4701,8 | 4,8 | 13,6 | 15,3 |
| Vascular system | 887 | 3868,1 | 4,4 | 12,4 | 12,6 |
| defined comorbidities | 703 | 2922,9 | 4,2 | 9,8 | 9,5 |
| Heart valve | 681 | 2600,2 | 3,8 | 9,5 | 8,5 |
| invasive therapy | 615 | 2149,7 | 3,5 | 8,6 | 7,0 |
| laboratory/technology/documentation | 518 | 2258,1 | 4,4 | 7,2 | 7,4 |
| Other comorbidities | 300 | 1474,4 | 4,9 | 4,2 | 4,8 |
| Total | 7156 | 30661,8 |  | 100,0 | 100,0 |

Defined comorbidities, which also included literature on COVID, achieved medium relevance with 9.8% of publications and an average of 4.2 IF per publication. Literature on COVID accounted for 5.16% of publications in 2020 and 2022, comparable to TAVI/TAVR, arrhythmias and heart valves over the entire observation period.

### Study Types

The study types and associated IFs are listed in Table 4. Retrospective studies (19.8% of all publications), basic research on cells and animals (13.4%), and (systematic) reviews (13.0%) were the most common study types. They accounted for 18.9%, 18.3% and 14.5% of all c-IFs, respectively. The highest average IF per individual publication was achieved in randomized controlled trials (mean IF 7.5), basic research on cells and animals (IF 5.8), laboratory studies (IF 5.4), and meta-analyses (IF 5.4). The lowest values were found in case reports (IF 1.4) and review articles (IF 1.7).

**Table 4:**
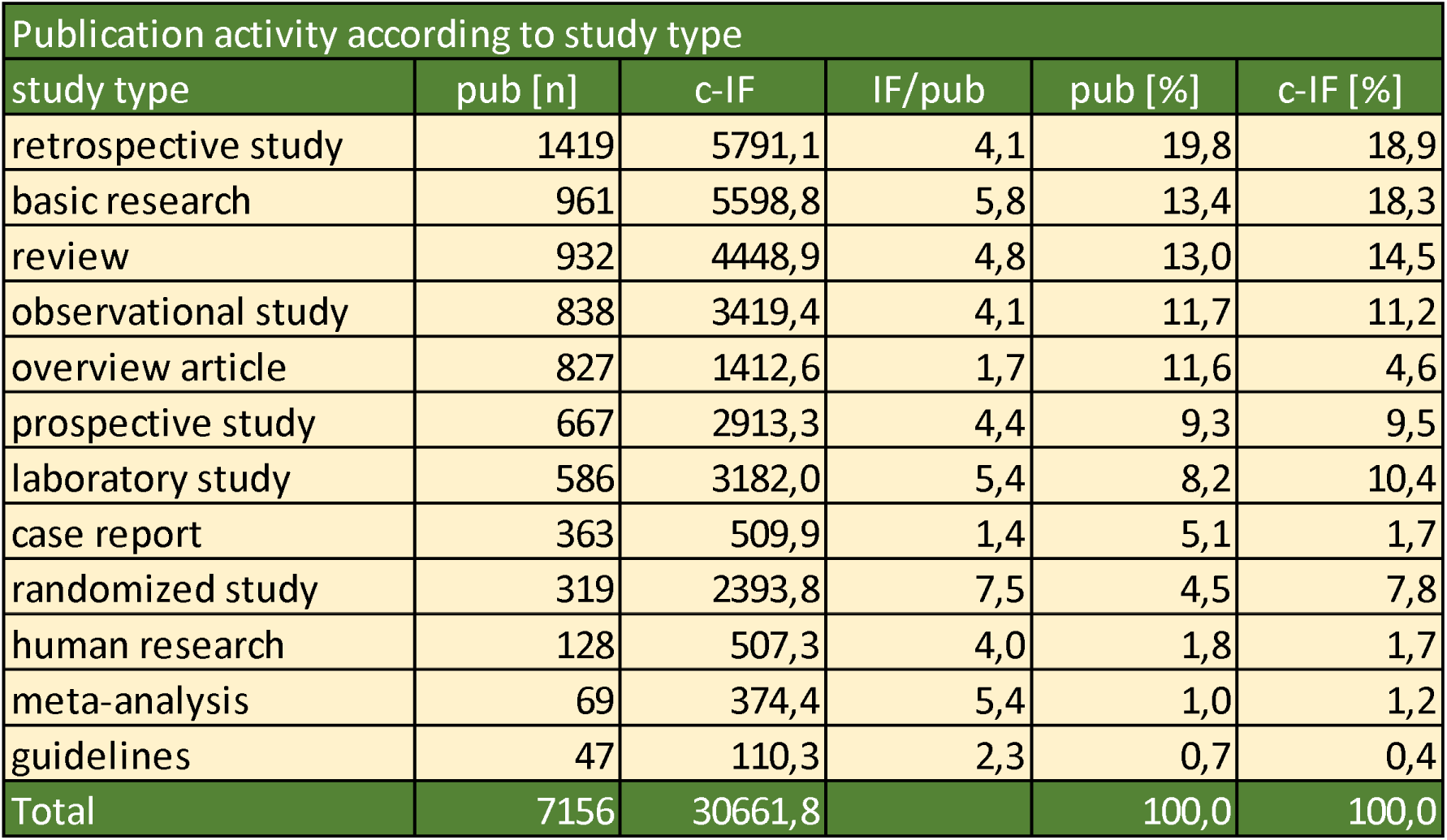
Publication activity according to study type in 39 cardiology departments based on the total number of publications (n=7156). All double authorships were excluded from this table. c-IF = cumulative impact factors; pub = publications

### Publications by Hierarchical Position and Gender of Staff

Of the total 672 employees, 530 (78.9%) were male and 142 (21.1%) were female. The proportion of female employees decreased with increasing hierarchical level (directors: 0/48; senior consultants: 37/210 (17.6%); consultants: 105/414 (25.4%)) and with increasing academic degree (professors: 23/215 (10.7%); assistant professors (private lecturers; PD): 41/154 (26.6%); doctors with a PhD: 70/274 (25.5%); without academic title: 8/29 (27.6%).

The number of publications and the resulting c-IF, categorized by academic title and position in the department (director/advisor) and differentiated by gender, are shown in Table 5. A clear hierarchical gradient was observed in the number of publications and the c-IF. Professors had the highest number of publications per employee (men: 27.8; women: 15.8), followed by assistant professors (PDs) with 11.7 (men) and 8.9 (women) publications per employee, respectively. Professors also achieved the highest c-IF per employee. In line with their academic title, directors had a significantly higher number of publications per employee (men: 38.8; women: 0/0) than senior physicians (men: 16.6; women: 9.3). Among the directors, 1,566 of 1,861 publications (84.1%) were last authors; among senior consultants, 2,125 of 3,210 (66.2%) were last authors, and among consultants, 1,497 of 3,208 (46.7%) were last authors. Among professors, 4,217 of 5,710 (73.9%) last authors, compared to 707 out of 1,682 (42.0%) among assistant professors, 230 out of 844 (27.3%) among physicians, and 34 out of 43 (79.1%) among consultants without academic titles (see Figure 1).

**Figure 1:**
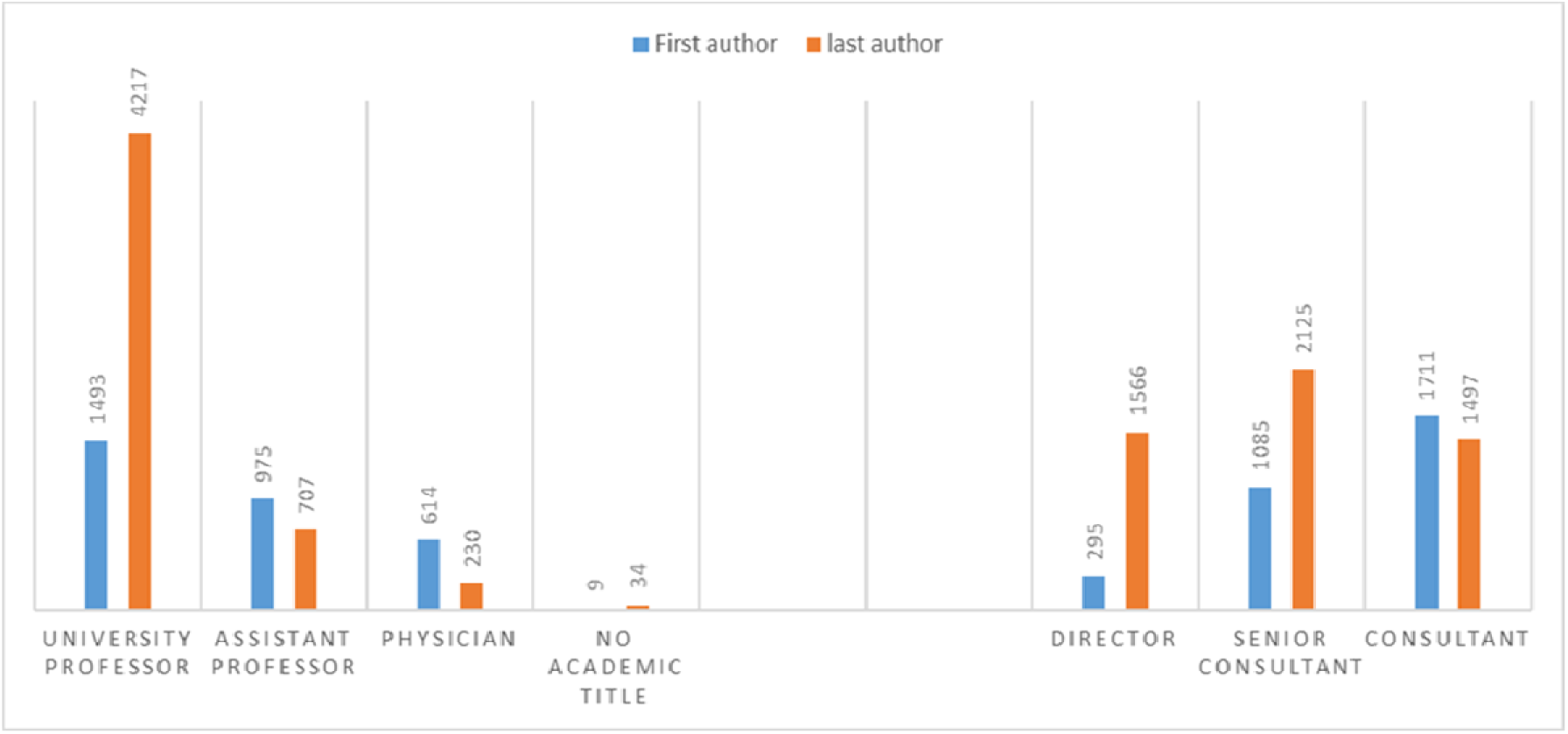
The figure shows the ratio of first and last authorship within academic levels and between hierarchical levels at university hospitals.

**Table 5:** Publication activities according to author hierarchy and taking into account the gender of the respective hierarchical level. All first and last authorships were taken into account in this table; the number of authorships (n = 8279) is therefore higher than the number of pub-lications (n = 7156). c-IF = cumulative impact factors

| Publication activity by author hierarchy |  |  |  |  |  |  |  |
| --- | --- | --- | --- | --- | --- | --- | --- |
| Degree /<br>Hierarchical position | Staff |  |  | Publication/gender |  | c-IF/gender |  |
|  | total | Female | Male | Female | Male | Female | Male |
| Professor | 215 | 23 | 192 | 15,8 | 27,8 | 67,4 | 125,7 |
| Assistant professor | 154 | 41 | 113 | 8,9 | 11,7 | 29,7 | 45,4 |
| Doctor | 274 | 70 | 204 | 2,4 | 3,3 | 7,4 | 11,3 |
| No academic title | 29 | 8 | 21 | 0,6 | 1,8 | 2,5 | 7,1 |
| Director | 48 | 0 | 48 | 0,0 | 38,8 | 0,0 | 197,8 |
| Senior consultant | 210 | 37 | 173 | 9,3 | 16,6 | 36,8 | 65,9 |
| Consultant | 414 | 105 | 309 | 5,3 | 8,6 | 19,0 | 34,7 |

Publication output by gender is shown in Table 6. Of the 672 employees, 538 (80.1%) had published at least once. Among men, 82.5% (437/530) had published, compared to 71.1% (101/142) of women. Women accounted for 21.1% of the total workforce and 18.8% of publishing employees. On average, male authors had 13.9 publications per person, while female authors had 6.3. The total c-IF (35,067.0) was distributed 90.5% to male employees and 9.5% to female employees. The average IF per publication was 4.3 for men and 3.7 for women. Further details are shown in Table 6.

**Table 6:** Publication activities of male and female authors. All first and last authorships were taken into account in this table; the number of authorships (n = 8279) is therefore higher than the number of publications (n = 7156). c-IF = cumulative impact factors

| Publication activity by gender of authors |  |  |  |  |  |  |
| --- | --- | --- | --- | --- | --- | --- |
|  | Male | Male [%] | Female | Female [%] | total | total [%] |
| Employees (n) | 530 | 78,9 | 142 | 21,1 | 672 | - |
| publishing employees (n) | 437 | 82,5 | 101 | 71,1 | 538 | 80,1 |
| First authorship (n) | 2574 | 83,3 | 517 | 16,7 | 3091 | 37,3 |
| Last authorship (n) | 4804 | 92,6 | 384 | 7,4 | 5188 | 62,7 |
| Authorships in total | 7378 | 89,1 | 901 | 10,9 | 8279 | - |
| Authorship / Employees | 13,9 | - | 6,3 | - | 12,3 | - |
| Authorship / pub. Employee | 16,9 | - | 8,9 | - | 15,4 | - |
| c-IF First authorship | 12196,3 | 85,8 | 2022,9 | 14,2 | 14219,2 | 40,5 |
| c-IF Last authorship | 19549,1 | 93,8 | 1298,6 | 6,2 | 20847,8 | 59,5 |
| c-IF | 31745,4 | 90,5 | 3321,6 | 9,5 | 35067,0 | - |
| IF per authorship | 4,3 | - | 3,7 | - | 4,2 | - |

## Discussion

The aim of this bibliometric analysis was to evaluate the publication output of leading staff members in German university cardiology departments over a period of 10 years. The analysis focused specifically on first and last authors, as had previously been done for surgical specialties [3-5]. In addition to the number of publications, the study also aimed to describe the topics and focus of the publications as well as the type of studies conducted. The publications were evaluated using the impact factor, a controversial quality indicator. Although this is considered a quality benchmark for journals, it is often mistakenly used as a quality indicator for individual articles within these journals. Nevertheless, it is a suitable tool for classifying the overall publication performance of clinical departments within the same specialty, as confirmed by the similar results of a comparative study using the journal-based h-index [4].

Since this is the first study to examine the publication output of cardiology departments at German universities—and no comparable study for other countries was found in a PubMed search—the data presented can only be compared to a limited extent. The study represents a snapshot of the publication activity of cardiology departments at German universities over a decade. This allows it to form the basis for analyzing progress in cardiology research. By analyzing the gender distribution at the management level, it also offers the opportunity to assess the necessary promotion of female physicians and researchers in university cardiology management, as desired by the DGK (DGK Annual Report 2022) [22].

Cardiology topics were grouped under the following specialist areas: Cardiology (45.14%) and interventional cardiology (32.29%) accounted for the majority of publications (over 77%), with almost all university hospitals (38 out of 39) publishing articles on these topics. Almost all university hospitals published on the specialist areas of “non-invasive diagnostic procedures” (37) and intensive care/emergency medicine (32). Twenty-six hospitals published on angiology (peripheral). Only 12 and 7 university hospitals published on neurology and pneumology, respectively (103 of 7156 publications; 1.44%). This reflects the variable, interdisciplinary structure of cardiology at German university hospitals. Depending on the university hospital, the specialist areas are combined in different ways or represent independent departments of the university institution.

This study identified coronary heart disease, cardiac arrhythmias, and heart failure as the main focus of German cardiological research. This largely corresponds to the main topics of German cardiological research identified by Millenaar et al. [1]: coronary heart disease, cardiac arrhythmias, and heart valve disease, followed by heart failure. Chorro et al. [6] identified TAVI (transcatheter aortic valve implantation), cardiac resynchronization therapy, and oral anticoagulation with non-vitamin K antagonists as important publication topics in Spain. Bouleti et al. [7] cited interventional cardiology, electrophysiology, cardiovascular imaging, and heart failure as the main topics in France. However, a detailed analysis comparable to that presented in Tables 1–4 has not yet been published. In addition, the present analysis distinguished between study type and specific areas of focus such as outcomes, diagnostics, and therapy. Furthermore, the objective of Millenaar et al. [1] to analyze cardiological research as such differed fundamentally from the present study, which, for reasons of comparability, deliberately focused on the publication activity of clinically active university cardiologists in leadership positions and included research laboratory staff in management positions who were listed on the website as part of cardiology.

The publication activities in cardiology described here can be directly compared with those in visceral and trauma surgery using the same methodological approach. Böckmann et al. [3] reported—also over a period of 10 years—on the publication activities of 442 university physicians and senior physicians in visceral surgery in 38 German departments, while Preut et al. [4] examined the publication output of 523 employees in 39 German university departments for trauma and orthopedic surgery. Visceral surgery produced 4,699 publications, which corresponds to 10.6 publications per employee; trauma surgery had 4,267 publications, or 8.2 publications per employee. This study found 10.6 publications per employee in cardiology (7,156 publications among 672 people), thus reflecting a comparable output of cardiology and visceral surgery in terms of average publication output per person.

In addition, the proportion of publishing employees in cardiology was 80.1%, which was higher than in trauma surgery (72.8%) but similar to visceral surgery (80.8%). The main difference between the surgical specialties and cardiology was in the number of cumulative impact factors (c-IF) generated. In visceral surgery, 4,699 publications achieved a total of 14,130.23 c-IF, which corresponds to an average of 3.0 IF per publication. Trauma/orthopedic surgery achieved 7,726.5 c-IF from 4,267 publications, corresponding to an average of 1.81 IF per publication. In contrast, cardiology achieved 30,661.8 c-IF from 7,156 publications – an average of 4.3 IF per publication.

Schwarzer et al. [8] had already determined that the IF per publication in university cardiology is higher than in general and cardiac surgery, which they attributed to the larger number of employees. However, since the number of publications per employee is identical in cardiology and visceral surgery, this explanation alone is not sufficient. Rather, this discrepancy underscores the fact that the IF is strongly influenced by the field of research [9]. Basic science studies—such as research with cell and animal models or laboratory studies—achieved significantly higher IFs in the present analysis than clinical (retrospective) observational studies and are more common in cardiology than in surgical specialties (see Table 4). In addition, collaborative projects in cardiology, as mentioned by Millenaar et al. [1], may also have contributed to the higher IF per publication in this field.

A particular focus of this study was the analysis of publication activity with regard to the hierarchical level of the authors and the gender of the employees. Since longer professional experience, generally leads to a higher hierarchical level, the expected differences in the number of publications between the various hierarchical levels were identified. The number of publications per employee decreased continuously from professors to assistant professors and physicians to employees without titles (Table 5), and the same was true for the comparison between directors and senior physicians. Female employees authored 10.9% of all first and last authorships, which is a higher proportion than in visceral surgery (8.7%) [10] and trauma surgery (6.1%) [4] during the same period. The proportion of women was 16.7% among first authors and 7.4% among last authors, compared with 11.0% female first authors and 7.0% last authors in visceral surgery [10] and 8.0% and 4.3% in trauma surgery and orthopedics [4], respectively. Women in leadership positions in cardiology were therefore slightly better represented in the author data than in the surgical specialties. However, the proportion of women as first and last authors was significantly lower than the figures reported by Millenaar et al. [2] for global cardiology publication activities. These authors reported that of a total of 257,940 articles, 35.4% of first authors and 22.9% of last authors were women. Amankwah et al. [11] also reported a higher proportion of women than observed here in their analysis of publications from 2016 in the American Journal of Cardiology, with 23% of first authors and 20% of last authors being women. Asghar et al. [12] found that 20.8% of first authors and 12.3% of last authors in 11,529 publications in six leading cardiology journals in 2016 were women. In another study on publication activities in cardiology over a period of 40 years (55,085 articles), Ouyang et al. [13] found that 26.7% of first authors and 19.7% of senior authors were women, emphasizing that women continue to be significantly underrepresented as senior authors, as was also the case in the present study. Petrechko et al. [14] came to a similar conclusion when analyzing 31,540 articles in the 20 most frequently cited cardiology journals. In 27% of the articles, women were the first authors and in 20% they were senior authors, while only 23% of the editorial board members were women. The significantly lower publication activity of female employees found here compared to the studies cited can most likely be explained by the methodology, as only the publications of clinically leading employees were recorded, but not those of younger employees.

On the other hand, there are considerable geographical differences in publication activities. In their analysis of 18,535 publications from leading cardiology journals (Journal of the American College of Cardiology, European Heart Journal, Journal of the American Medical Association Cardiology, Nature Reviews Cardiology), Goel et al. [15] reported that 20.6% of authors were women. In publications from the US, 34.6% of first authors were women, followed by 25.2% in publications from the UK, but only 4.9% in publications from Germany. The proportion of women as last authors also varied considerably by geography, with 47.6% for publications from the US and only 6.3% and 4.4% for publications from the UK and Germany, respectively. An analysis by Whitelaw et al. [16] also showed that women continue to be significantly underrepresented in randomized trials on heart failure.

In the present study, the proportion of female employees among senior staff in cardiology was 21.1%, among senior physicians 25.4%, but among senior physicians with management functions only 17.6%, and there were no female clinic directors. The proportion of women in leadership positions in clinical areas of German university medicine was documented by the German Medical Women’s Association [17]. For the year 2024, they reported a proportion of women of 11% among department heads in internal medicine. Blumenthal et al. [18] reported on the proportion of women among academic cardiologists in the USA. In 2014, 630 (16.5%) of 3,810 cardiologists with a university position were women. Women had fewer publications than men (16.5 vs. 25.2 on average; p < 0.001) and were less likely to hold a full professorship (30.6% of men vs. 15.9% of women). It was particularly emphasized that, when comparing eight specialties in internal medicine, women had lower chances of becoming full professors than men only in the fields of infectious diseases and cardiology; in the other six specialties of internal medicine, there were no gender-specific differences. No clear explanation was given for this last observation. A gender imbalance in academic positions in cardiology was also found in an analysis of 17 Canadian universities [19]. Of 1,040 cardiologists, 80.4% were men. The higher the academic position, the lower the proportion of women: 13.5% among full professors, 17.4% among associate professors, and 26.3% among assistant professors.

The causes of the underrepresentation of women are manifold, and this paper can only speculate on the reasons for the lower proportion of women in publishing activities. The German Medical Women’s Association [17] lists possible reasons and asks, among other things, whether structural discrimination exists and whether job advertisements sufficiently address flexible working hours, comprehensive childcare, the emphasis on teamwork and communication skills, and proof of good teaching evaluations in order to appeal to women. However, traditional role models still play a major role in professional life, including, in particular, the high amount of time required for care work. In a 2020 survey, the Allensbach Institute for Public Opinion Research found that 73% of mothers see responsibility for raising and caring for children as their task; this figure has improved by only three percentage points since the 2010 survey [23]. In a study (Capranzano 2016), women stated that, among other things, a lack of opportunities, concerns about radiation exposure, and prejudice from their male colleagues had prevented them from choosing interventional cardiology [25]. The DKG presented a position paper and adopted measures to promote women (2022) [22]. Since the cut-off date for this study is 2021, this work can serve as a basis and reference for the effectiveness and implementation of measures to promote women. A study from the UK found that women make up 16% of cardiology specialists and 29% of doctors in cardiology training. Women’s interest in cardiology has not increased over the past two decades, even though the proportion of female medical students has risen. Interventional cardiology and electrophysiology in particular attract less interest from women than from men [20]. The author described the culture in cardiology as not female-friendly, citing as reasons the poor work-life balance due to the double burden of family and career, limited mentoring opportunities, and an insufficient supply of attractive positions for women. An anonymous survey of medical students in Australia found that an equal proportion of male and female medical students aspire to a career in cardiology, although the majority cite poor work-life balance as the main obstacle. The high proportion of women who are already interested in cardiology during their medical studies suggests that the obstacles that deter or discourage women only arise at a later stage of clinical training [21].

This study has limitations that should be considered when interpreting the results. The academic roles of the authors were assessed based on the websites of the university departments on September 1, 2021. This represents a snapshot at that particular point in time and does not reflect the performance of individual departments over the entire study period, but only the performance of current senior staff. This approach may lead to a discrepancy between the publication data and the actual composition of the team in earlier years of the observation period, especially in cases of staff turnover or institutional changes. Therefore, when interpreting long-term publication trends, it must be taken into account that not all of the individuals listed necessarily held the same position throughout the entire ten-year period. Furthermore, it cannot be guaranteed with certainty that the information on the websites always accurately reflects the actual rank or academic degree of the authors being evaluated. The academic degree held by the author on the cut-off date has not been verified. It is therefore possible that some individuals have been classified with a lower academic degree. This may lead to minor distortions in the data. It should be noted that there is a high number of last authorships in the group “without academic title” compared to “doctors,” which can be attributed to a single person (1 first and 32 last authorship). Publications by assistant physicians and assistant physicians in further training were not taken into account, as their relatively short professional careers would not be representative for a period of ten years. Furthermore, only first and last authors were taken into account in order to assign clear scientific responsibility and leadership roles. Although this methodological decision improves comparability and focus, it may underestimate the extent of collaborative research, especially in large multicenter studies.

Nevertheless, this study is the first to provide a comprehensive overview of the publication activity, focus areas, topics, and gender of authors who are senior staff members in German university cardiology departments. The study examined not only publications in leading cardiology journals, but also all first and last authors who are chief physicians and senior physicians in a total of 876 journals. This allows for comparison with other fields. Such studies should be conducted regularly to evaluate scientific performance and to determine how the increasing number of female applicants will affect publishing activity in the future—and how this activity can be further promoted.

## Data Availability

All data produced in the present work are contained in the manuscript

